# Role of IL-23 neutralization in psoriasis – insights from a mechanistic PK/PD model and meta-analysis of clinical data

**DOI:** 10.1101/2021.03.07.21253086

**Authors:** Georgi I. Kapitanov

## Abstract

Blocking of IL-23 has shown a profound effect on patient outcomes in psoriasis. The current IL-23 binding monoclonal antibodies show differences in dosing regimens, pharmacokinetics, affinity for the target, and efficacy outcomes in the clinic. The goal of the current work is to use a mechanistic pharmacokinetics/pharmacodynamics mathematical model to estimate projected free IL-23 neutralization for the different therapeutic molecules and connect it to clinical efficacy outcomes. The meta-analysis indicates a sigmoid-like relationship and suggests that the best current anti-IL23 antibodies are close to saturating the efficacy that can be achieved by this pathway in psoriasis.

## Introduction

Cytokines are soluble signaling proteins that participate in many inflammatory processes and can initiate and propagate a variety of inflammatory indications and autoimmune diseases – rheumatoid arthritis, atopic dermatitis, inflammatory bowel disease, and psoriasis, among others (1-4). Using therapeutic antibodies that bind directly to the cytokine or its receptor on immune cells to neutralize cytokine-mediated signaling, has shown to be an effective strategy for mitigating these diseases. Some examples include adalimumab (Humira) and infliximab (Remicade), anti-TNFα monoclonal antibodies (mAbs) that have been approved for treatment in psoriasis, rheumatoid arthritis (RA), and inflammatory bowel disease (IBD) (5); dupilumab (Dupixent), a mAb which modulates interleukin (IL-)4 and IL-13 signaling by blocking IL-4 receptor α (IL-4R α) and has been approved for atopic dermatitis and asthma (6, 7); secukinumab (Cosentyx), an anti-IL17A mAb approved for psoriasis (8); tocilizumab (Actemra), an anti-IL-6R mAb approved for RA (9).

Psoriasis is an auto-immune disease characterized by dry, scaly skin patches that affects over 100 million people worldwide. Common co-morbidities are diabetes, heart disease and depression (10). Modulating the Th17 pathway has had a profound effect on mediating the pathogenesis of psoriasis, both through blocking IL-23 and through blocking IL-17A (8, 11-14). Some of the current biologic therapeutics on the market achieve clear to almost clear skin in more than 70% of patients (15-18), which allows for a small margin of improvement for any future therapeutics.

IL-23 is a heterodimeric cytokine comprising two subunits, IL12p40 (shared with IL-12) and IL23p19. It is secreted by antigen-presenting cells and keratinocytes and plays a role in the proliferation of Th17 cells, which are implicated in chronic inflammation associated with psoriasis (8, 11-14). Ustekinumab was the first approved anti-IL-23 therapeutic, targeting the IL12p40 subunit of IL-23, hence blocking both IL-12 and IL-23 activity. Discoveries suggesting a key role for IL-23 in the pathophysiology of psoriasis led to follow on therapeutics that solely target the IL23p19 subunit of IL-23, which spares IL-12 signaling (19).

The current work attempts to shed light on the role of blocking IL-23 in psoriasis through a mechanistic model of the pharmacokinetic/pharmacodynamic (PK/PD) interaction between the IL-23 blockers and IL-23. The model quantifies the effect of projected IL-23 neutralization on clinical endpoints in psoriasis. This modeling effort has implications for the future development of IL-23 blockers in psoriasis: using PK data and affinity to IL-23, one can project the efficacy that would be observed in the clinic at a particular dosing regimen.

## Methods

### PK/PD model set up

A two-compartment model that incorporates drug:target binding interactions in the central (plasma) compartment was used. Interactions of anti-IL12p40 drugs (ustekinumab and briakinumab) with only IL-23 were considered; binding/unbinding to IL12p40 monomer and homodimer, as well as IL-12, were ignored. The model scheme can be seen in Figure 1 and the model equations are described in **Error! Reference source not found**., where *D*_*P*_, *T*, and *C* represent the concentrations in plasma of free drug, free IL-23, and drug-target complex, respectively *D*_*T*_. is the drug concentration in the peripheral tissue compartment and *D*_*D*_ is the drug in the subcutaneous depot compartment. *Dose*(*t*) is the dose, which is administered subcutaneously at a certain interval. Target synthesis and elimination are assumed only in the Central compartment.

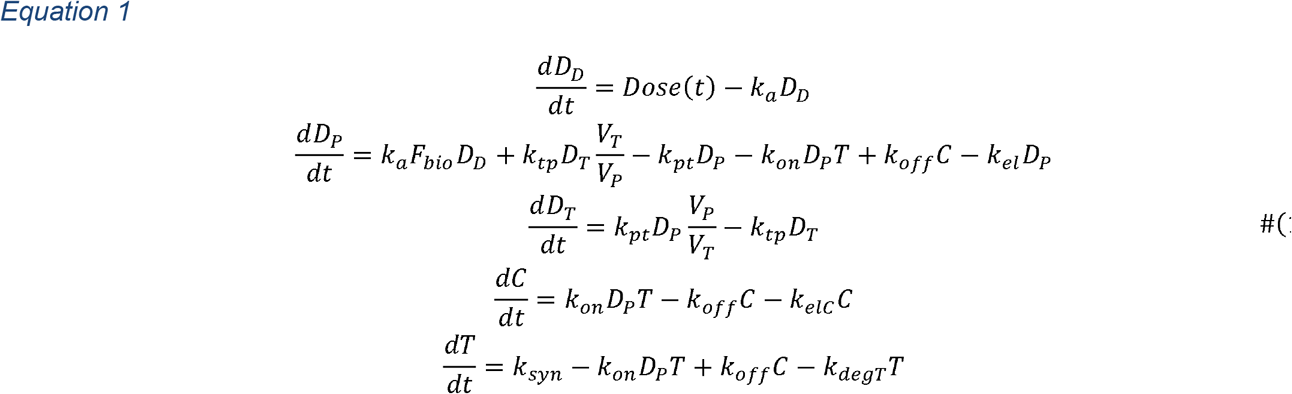

**Figure 1:**
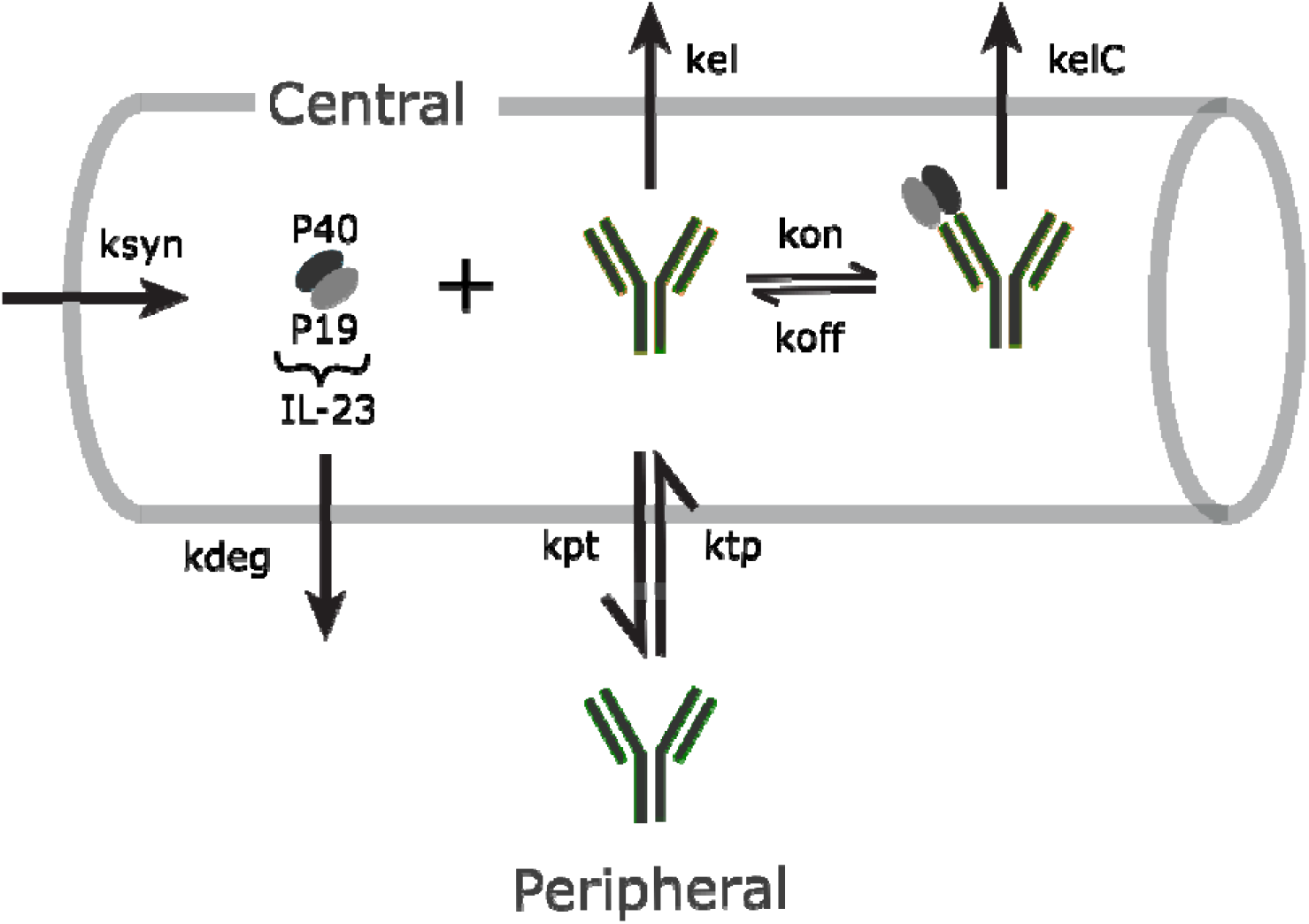
Model Scheme for anti-IL23 mAbs. After subcutaneous administration, the antibody is in the Central compartment (plasma) and can distribute back and forth to the Peripheral compartment. IL-23 is synthesized and degraded endogenously in the Central compartment only. Binding and unbinding of the mAb with IL-23 as well as the clearance of the drug:target complex are restricted to the Central compartment.

Table ITable I: Definitions and common values of parameters used in the model contains a description of the model parameters and common estimated values used in the model. Table II contains the binding affinity and pharmacokinetic parameters used for each drug.

**Table I:** Definitions and common values of parameters used in the model

| Parameter | Description | Value | Units |
| --- | --- | --- | --- |
|  | Central compartment volume | Drug specific | L |
|  | Drug distributive clearance rate | Drug specific | L/day |
|  | Drug elimination clearance rate | Drug specific | L/day |
|  | Peripheral tissue volume | Drug specific | L |
|  | Rate of absorption post subcutaneous drug administration | Drug specific | 1/day |
| $F_{bio}$ | Bioavailability post subcutaneous drug administration | Drug specific | dimensionless |
| $k_{on}$ | Drug-target concentration-dependent association rate | 86.4 (20) | $\text{nM}^{-1}\text{day}^{-1}$ |
| $K_D$ | Drug-target dissociation constant | Drug specific | nM |
| $k_{off}$ | First-order dissociation rate constant of antibody | $k_{on}K_D$ | 1/day |
| $k_{el}$ | Rate constant of drug elimination from plasma | $\frac{Cl}{V_P}$ | 1/day |
| $k_{pt}$ | Rate constant of drug distribution from plasma to peripheral tissue | $\frac{Q}{V_P}$ | 1/day |
| $k_{tp}$ | Rate constant of drug distribution from peripheral tissue to plasma | $\frac{Q}{V_T}$ | 1/day |
| $k_{degT}$ | Rate constant of target elimination from plasma | 4.16 (21) | 1/day |
| $k_{syn}$ | Zero order target synthesis rate in SoA | $k_{degT}T(0)$ | nM/day |
| $k_{elC}$ | Rate constant of complex elimination from plasma | $k_{el}$ | 1/day |

**Table II:** Modeling parameters for anti-IL-23 mAbs

| Drug<br>Parameter | Ustekinumab<br>(23) | Briakinumab<br>(24)* | Guselkumab<br>(25)* | Risankizumab<br>(26) | Tildrakizumab<br>(27)* |
| --- | --- | --- | --- | --- | --- |
| $V_p$, L | 3.0 | 3.0 (0.049) | 3.1 (0.098) | 5.7 | 2.3 (0.031) |
| $V_T$, L | 1.4 | 3.5 (1.3) | 3.9 (0.94) | 3.4 | 2.7 (0.12) |
| $Q$, L/day | 0.16 | 0.81 (0.14) | 0.89 (0.27) | 0.33 | 1.3 (0.074) |
| $Cl$, L/day | 0.19 | 0.74 (0.093) | 0.30 (0.047) | 0.30 | 0.15 (0.0044) |
| $k_a$, 1/day | 0.14 | 0.24 (0.086) | 0.31 (0.23) | 0.18 | 0.30 (0.059) |
| $F_{bio}$, % | 87 | 41 (8.8) | 42 (16) | 72 | 79 (7.3) |
| $K_D$, pM | 120 (28) | 100 (24) | 3 ** | 1 (29) | 300 (30) |

\* Mean reported data were digitized and fit to a two-compartment PK model. Mean parameter fit results are reported and used in the modeling, standard errors in parenthesis). Average human bodyweight assumed to be 70 kg. Fits were performed using MATLAB Simbiology.
\*\* FDA BLA document

Target neutralization was calculated as *TC* = 1 – *T*/*T*(0). The average target neutralization at a time point *t*_*p*_ was calculated as 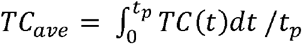 or, similarly to standard PK notation, target neutralization 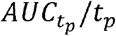. *T*(0) is 0.7 pM (22) and IL-23 half-life is fixed at 4 h (21).

### Data collection and analysis

The efficacy endpoints chosen were % of patients who achieved a certain % reduction in Psoriasis Area and Severity Index (i.e. PASI 75 refers to % of patients who achieved a 75% reduction in their PASI score). Data from available clinical trials was collated, efficacy endpoints (PASI 75, PASI 90, and PASI 100) were collected where available, and number of patients in each trial and cohort noted. Clinical data combined psoriasis and plaque psoriasis data and comprises four studies for ustekinumab (Phase (Ph) 2 (31) and Ph3 Accept (32) and Phoenix 1 & 2 (33, 34)), two studies for briakinumab (Ph2 (35) and Ph3 (36, 37)), five studies for guselkumab (Ph1 (38), Ph2 (39, 40), Ph3 VOYAGE 1 & 2 (18, 39), and Ph3 ECLIPSE (41)) three studies for risankizumab (Ph2 (29) and Ph3 ULTIMMA 1 & 2 (15), ustekinumab data from these studies were excluded because it was not dose-separated and results were consistent with already available data), and three studies for tildrakizumab (Ph2b (30) and Ph3 reSURFACE 1 & 2 (42)).

The final efficacy readout used in the model was done on an intent-to-treat basis and placebo corrected. If a placebo group was switched to drug administration, they were assumed to be a different cohort and their target neutralization estimated separately. If there was no available placebo data to correct efficacy at a particular time point or study, the nearest time point placebo efficacy read out was assumed to carry forward or the placebo efficacy read out from an equivalent phase study with the same drug was used.

Based on dosing regimens, average IL-23 neutralization was projected at the same time points at which efficacy data was collated. A visual description with an example can be seen in Figure 2. All efficacy results, the associated IL-23 neutralization, and the number of patients were pooled. A sigmoid curve with domain 0 to 1 was fit with IL-23 neutralization as independent variable and efficacy as dependent variable (Equation 2), each point in the meta-analysis weighted by the number of patients in the cohort from which the endpoint was collected. All model construction and fits were done in MATLAB.

**Figure 2:**
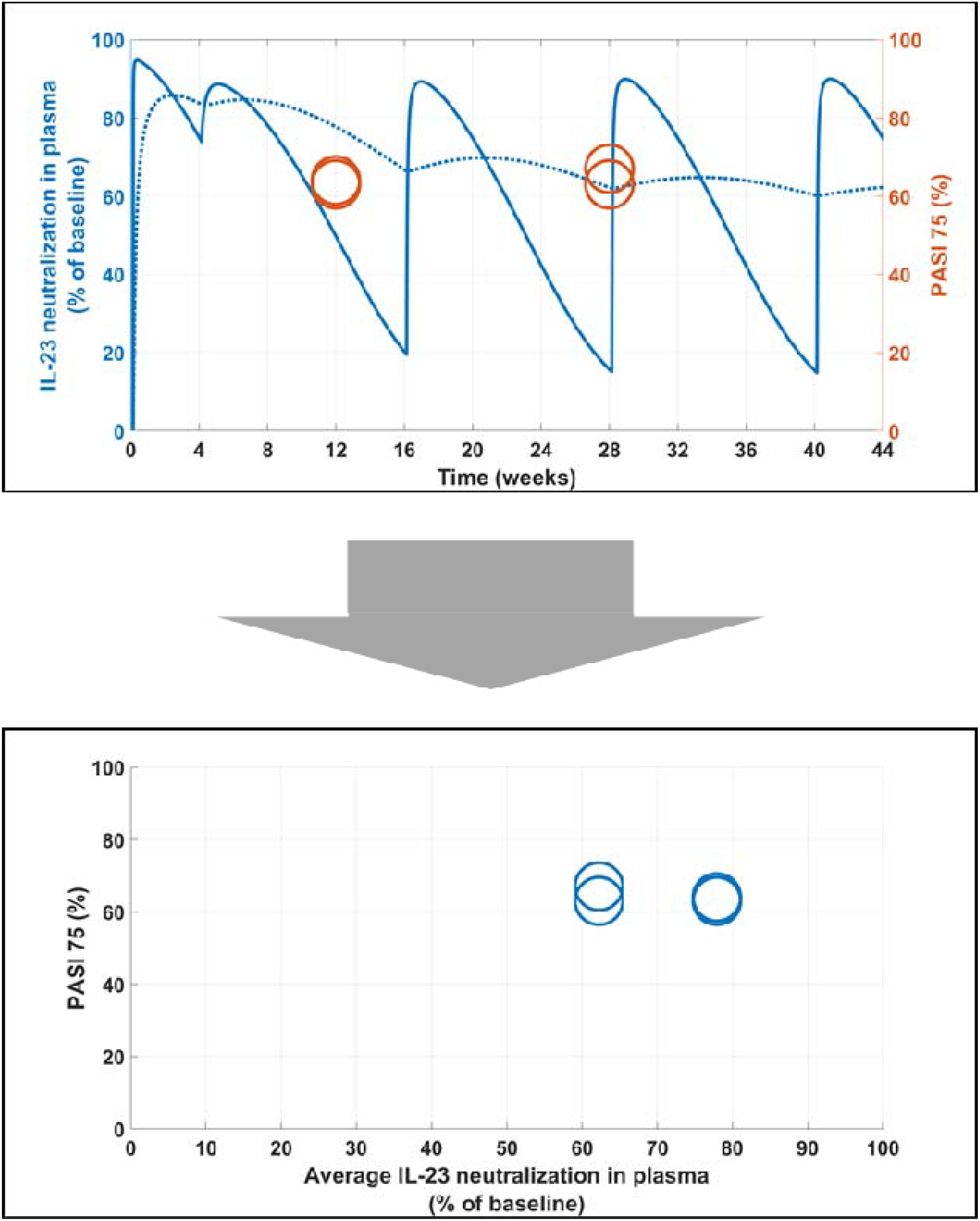
Steps in preparing the data for analysis. Step 1 (top): Based on PK and KD data for the molecule and dosing regimen in study cohorts, a prediction of IL-23 neutralization () is generated (solid blue line). Average target neutralization () is calculated from the prediction (dotted blue line). Efficacy endpoint score at particular time points are collated (open orange circles). The meta-analysis is based on projected average IL-23 neutralization in plasma and efficacy endpoints (bottom figure). The example presented is from ustekinumab Phase 3 PHOENIX 1&2 studies, where the dosing regimen is 45 mg subcutaneously every four weeks for two doses and every twelve weeks afterwards. PASI 75 endpoints were collected at week 12 and week 28. Two circles are shown at each graph – one for PHOENIX 1 and one for PHOENIX 2 results.

*Equation 2: is PASI 75/90/100, is the maximum efficacy achieved at 100% suppression of IL-23, is an inflexion-related parameter, TN is the projected IL-23 target neutralization in %, and h is a Hill-like power coefficient*.

## Results

The parameter value for *E*_x_ was consistently fit to 1 for all efficacy endpoints, therefore is fixed at 1. This rewrites Equation 2 as *E*= *E*_*max*_ * 2 * (*TN*/100)^*h*)/((*TN*/100)^*h* + 1), which reduces the fitting to parameters *h* and *E*_*max*_.

The model captures a trend of increased efficacy with projected increase of IL-23 neutralization but does not fully account for the variability in the efficacy results. The parameter estimates are in Table III. Given the *E*_*max*_ values, with full neutralization of IL-23 one would expect 78% of patients to experience 75% reduction in PASI score, 67% of patients to experience 90% reduction, and 42% to achieve full reduction of their PASI score. From the equation for *E*, one can derive a value for *TN*50, i.e. the target neutralization of IL-23 at which 50% of the maximum efficacy endpoint is projected to be achieved. With *E*_*x*_ fixed at 1, 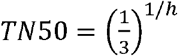. The TN50 values are reported in Table IV. For example, *TN*50 PASI 100 is 76%, which indicates that in order to achieve PASI 100 of 21% (half of *E*_*max*_ PASI 100), an IL-s23 blocker would need to neutralize 76% of plasma IL-23.

**Table III:**
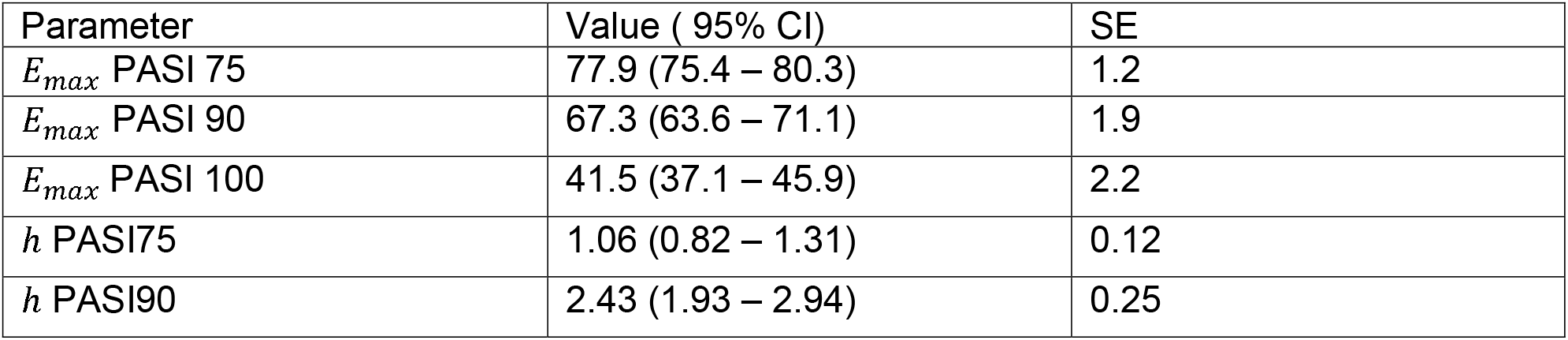
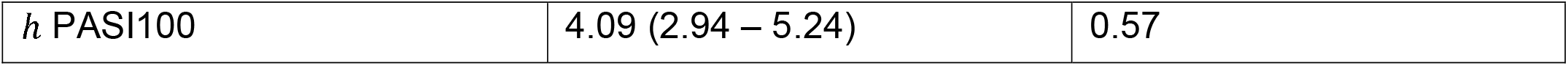
Parameter fit results

**Table IV:** TN50 calculations

| Parameter | Value |
| --- | --- |
| TN50 PASI75 | 35.5 % |
| TN50 PASI90 | 63.6 % |
| TN50 PASI100 | 76.4 % |

The confidence intervals around *E*_*max*_ increase with the PASI reduction needed to be achieved, which is expected since there would be many intra-patient factors that may affect the appearance of full or completely full clearance of skin and that would increase the variability of outcome.

The *TN*50 values also increase with PASI score reduction, which is also expected – it is harder to achieve PASI 90 than PASI 75, for example. This also leads to saturation of the efficacy responses at lower IL-23 neutralization for lower level of efficacy response. A visual check on Figure 3 A shows that ∼95% IL-23 neutralization saturates the PASI 75 response rate, B shows that ∼99% IL-23 neutralization saturates the PASI 90 response rate, and C indicates that ∼ 99.5% IL-23 neutralization saturates the PASI 100 response rate. Therefore, in order to differentiate from competitors, a new molecule needs to show efficacy at the harder to achieve endpoint. This conclusion is corroborated by the difference between earlier clinical trials where PASI 75 was enough to show differentiation (32, 40) and the more recent head-to-head trials where PASI 90 has become a primary endpoint (16, 39, 41).

**Figure 3:**
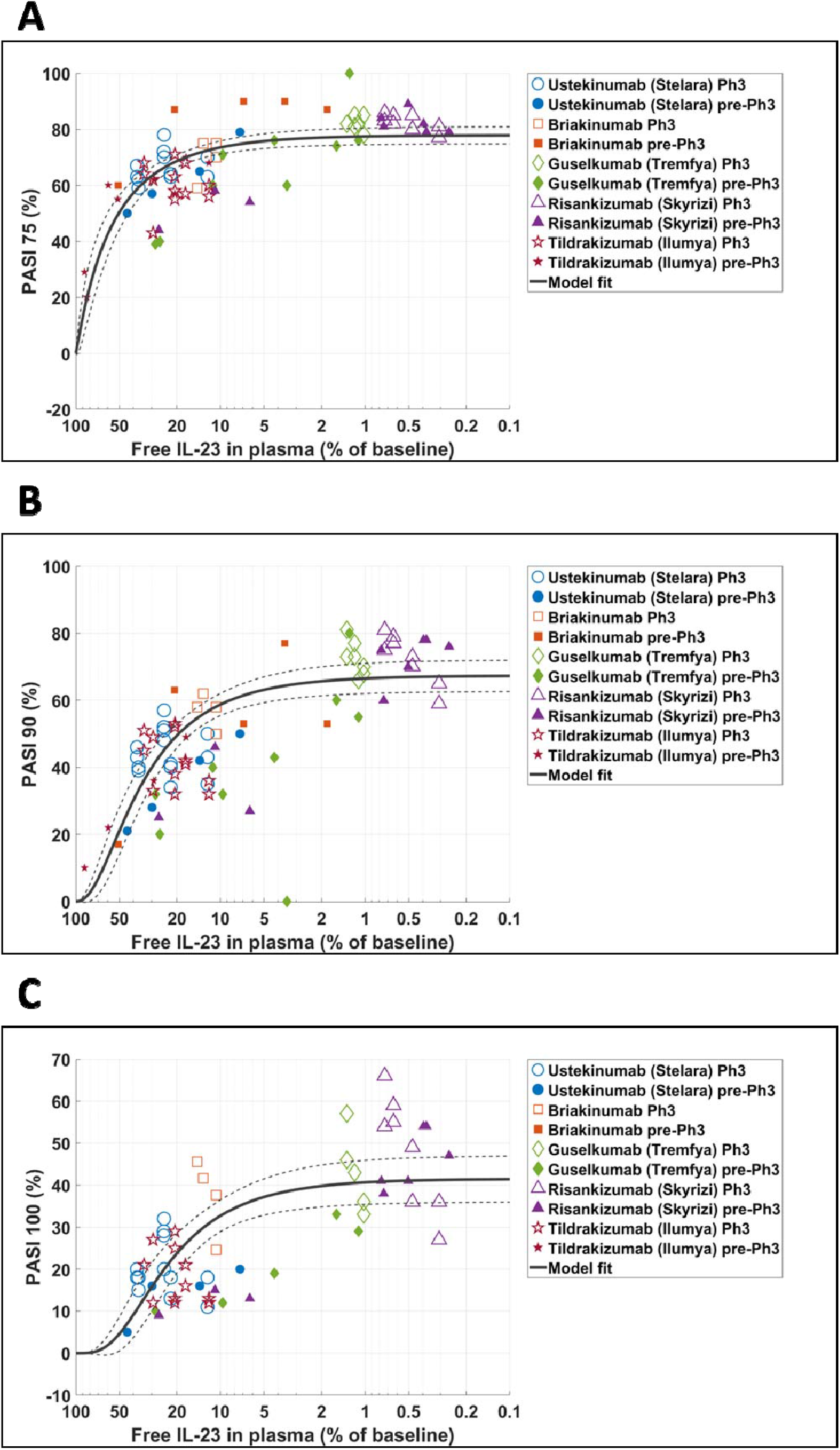
A visual representation of the fit of Equation 2 to the collated data and projected IL-23 neutralization. For presentation purposes, free remaining IL-23 is used on the x-axis. Projected free average IL-23 as % of baseline at different time points after drug administration is related to the average PASI 75 (A), 90 (B), and 100 (C) endpoints reported by the denoted drug at the same time point and the same doses using the function outlined in Equation 2. Open symbols denote data from Phase 3 trials, all others are from Phase 1 and 2 trials – the difference is used to indicate the weight of the points in the overall fit of the data. The percentage of free IL-23 was calculated as 100 times the ratio of simulated free IL-23 after drug administration and baseline IL-23. Results from the fits are reported in Table III.

## Discussion

Neutralization of IL-23 has proven to be an effective method for the treatment of psoriasis. Based on the analysis presented here, the greater the neutralization of IL-23, the better the efficacy. Ustekinumab (Stelara) was approved by the FDA in 2009, tildrakizumab (Ilumya) in 2018, guselkumab (Tremfya) in 2017, and risankizumab (Skyrizi) in 2019. The latter two show a significant increase in affinity to IL-23 through binding to the IL23p19 unit and achieving significant improvement in efficacy over the other competitors in the class. At the time of preparation of this manuscript, another anti-IL23p19, mirikizumab, announced promising results in psoriasis (43), which were followed up with an announcement that establishes it as a highly competitive molecule (44). At the time, no full article, including pharmacokinetics data, had been published to the author’s knowledge, hence mirikizumab data was omitted from the current analysis.

Similar comparative analysis of IL-23 neutralization among competitors was also done by Zhang et. al. (21). There are minor differences in drug:target interaction modeling between their approach and the present work. The goal in (21) was two-fold – to present results from recombinant IL-23 pharmacokinetics and to rank the molecules based on projected target neutralization in comparison to achieved PASI 100. Here the goal is to establish a general relationship between IL-23 neutralization and efficacy.

By targeting the IL12p40 subunit of IL-23, ustekinumab and briakinumab also target IL-12. Given the relatively low concentrations of cytokines in plasma, not explicitly accounting for the competition between IL-12 and IL-23 for binding to these therapeutics is reasonable. The method of analysis in the present work is not fit to statistically determine to what extent neutralizing IL-12 on top of IL-23 affects the efficacy read out for the two mAbs. However, their results are consistent with the overall behavior when combined with the rest of the available data. Therefore, based on the analysis conducted here, the efficacy achieved by both molecules appears due to blocking IL-23 only. Originally, ustekinumab and briakinumab were in development based on evidence for IL-12’s role in psoriasis (19). Curiously, IL-23 was discovered right around the time ustekinumab’s Investigational New Drug application with the Food and Drug Administration (FDA) was filed (FDA 75 FR 75678) and the therapeutic was being prepared for clinical trials (19). Further development of therapeutics targeting IL-23 without interacting with IL-12, by blocking the IL23p19 subunit, indicates that in practice IL-12 does not contribute significantly to the mitigation of psoriasis. Naturally, this hypothesis would have to be evaluated by clinical trials of IL-12 blockers that do not interact with IL-23, which, given clinical and pre-clinical data and the current state of development of therapeutics for the disease, is unlikely (19). This is partially due to the effectiveness of anti-IL23p19 antibodies, as well as to the safety outcomes of briakinumab (45). Although Stelara is considered a highly safe therapeutic (46, 47), deaths in briakinumab clinical trials have pinpointed excessive blocking of IL12p40 as a potential concern, evidence for which is underwhelming and may be related to other factors (45). Furthermore, ustekinumab has been approved for Crohn’s disease and ulcerative colitis, where blocking IL-12 along with IL-23 may have a more robust therapeutic role. More data from upcoming anti-IL23p19 blockers in IBD indications may shed more light on the role of each pathway in clinical efficacy.

Several points regarding the analysis performed here can be discussed further. The analysis is based on a 70 kg average patient, which is lower than the typical psoriasis patient. However, given that several compounds were analyzed and some of the PK parameters were taken from analysis in healthy volunteers as well as patients, it is unlikely that adding a bodyweight-based analysis would alter the overall results significantly. Early (induction), mid (around 6 months) and late (around 1 year) time points and projected IL-23 neutralization were analyzed together. Analysis was done with later time points removed and results did not differ substantially (data not shown), hence analysis including all time points is presented.

More variability was observed with the PASI 100 efficacy endpoint. The reasons may be several – 100% clear skin may be a more variable endpoint versus estimates of 90% or 75% improvement. Also, the variability at saturation comes from maintenance data from risankizumab and disappears when excluding late time points (results not shown). This result may be due to disease progression and another method of evaluation of IL-23 blockade that accounts for this may be a more robust predictor of efficacy.

While not the goal here, it would also be interesting to relate IL-23 neutralization to PASI change longitudinally. The longitudinal PASI data is sparse and not often presented in the literature. However, it is worth noting that an autoimmune disease like psoriasis can be effectively mitigated with a maintenance dosing regimen of every 8-12 weeks (48-51) versus another skin condition like atopic dermatitis, which appears to require a more frequent dosing of 2-4 weeks (52). This disparity may have implications for the target neutralization - efficacy relationship. On the other hand, while not providing superior efficacy, IL-23 blockers provide significantly improved dosing regimen compared to anti-IL17 blockers (53), so the difference in dosing regimens may be target-dependent as well. Analysis like the one done here can be performed to compare the anti-IL-17 blockers and propose a model-derived hypothesis for the difference in regimens.

The analysis conducted here also indicates that there is little room for improvement in outcomes for psoriasis patients with further neutralization of IL-23, given the saturating efficacy presented in Figure 3. However, the high level of IL-23 neutralization presented is partially the result of the target related parameters. Given the nature of interleukins and the consistently low protein levels detected in human plasma, it is unlikely that the baseline level of IL-23 would differ significantly from the concentration used here. However, the reference used to inform the rate of

IL-23 elimination is derived from cynomolgus monkey recombinant IL-23 PK study (21). All these factors may affect the endogenous turnover rate of IL-23 in humans. Given the already projected overall high neutralization, it is unlikely that the turnover in humans would be slower – this would only make the projected neutralization even higher and differentiation between the different molecules smaller, hence indicating that small differences in neutralization of the target lead to large differences in efficacy, the probability of which is low. However, higher turnover would mathematically result in faster IL-23 synthesis, which would decrease the level of neutralization. If significant, this would imply that there is further room for improvement in efficacy by increased blocking of IL-23. Given the reported very high affinities for risankizumab and guselkumab, the way to achieve this would likely be through applying even higher doses in psoriasis trials. Given the high efficacy of those two molecules at the current doses, this is unlikely to be tested in any robust manner in the near future.

The value of conducting modeling analyses like the currently presented one goes beyond comparison among molecules – for this purpose there are network meta-analyses of clinical data that utilize tools from pharmacometrics and statistics (53, 54). The model presented here can be used for informing future endeavors in the development of anti-IL-23 molecules for psoriasis but can also serve physicians for establishing a more robust confidence in superiority of one compound over another when head-to-head clinical data are lacking. Further, the model could also be used for projecting alternative dosing regimens for compounds for the purpose of improving efficacy better than a single compound exposure-response analysis because it includes a wider range of compounds and data. However, while the model can be potentially informative, real-world data would be needed to validate the projected efficacy and ensure the safety of the patients at these alternative regimens.

## Conclusion

A mechanistic model of free IL-23 serum neutralization and analysis of its effect on efficacy endpoints in psoriasis using available data from clinical trials was conducted. The results indicate that the higher the level of IL-23 neutralization, the better the efficacy and conclude, based on the high projected IL-23 neutralization, that there is little room for further improvement in the treatment of psoriasis through this pathway.

## Data Availability

The data is available in the literature and is referenced within the manuscript.

